# Regional Disruption of Slow-Wave Sleep Homeostasis in Children with Sleep-Disordered Breathing

**DOI:** 10.64898/2026.07.15.26358161

**Authors:** Gary Garcia Molina, Beth Peterson, Emma Strainis, Tony Kille, Annika Myers, Tâmara P. Taporoski, Camilla Matthews, Ana Maria Vascan, Stephanie Jones

## Abstract

**Importance:** Sleep-disordered breathing (SDB) is common in childhood and is associated with attentional and behavioral impairments despite largely preserved sleep macrostructure and minimal abnormalities in conventional electroencephalographic measures. This discrepancy has contributed to the perception that sleep is relatively preserved in pediatric SDB and has limited understanding of the physiological mechanisms underlying morbidity.

**Objective:** To determine whether pediatric SDB is associated with disruption of the regional organization and homeostatic dynamics of slow-wave activity (SWA), a key physiological marker of sleep-dependent neural recovery and development.

**Design, Setting, and Participants:** Cross-sectional study of 62 children aged 4 to 12 years who underwent overnight polysomnography with high-density electroencephalography in a laboratory setting. Participants were recruited from clinical referrals and the community, spanning the full spectrum of SDB severity.

**Exposures:** SDB severity indexed by hypopnea index (HI), apnea-hypopnea index (AHI), and obstructive apnea index (OAI).

**Main Outcomes and Measures:** Regional electroencephalogram-derived SWA (0.5–4 Hz) topography and exponential decay parameters derived from frontal and posterior cortical regions. The frontal-to-posterior decay-rate ratio was evaluated as a summary measure of regional sleep homeostasis.

**Results:** In children with lower hypopnea index, SWA demonstrated the expected developmental pattern, with posterior predominance in younger children and a progressive shift toward a more balanced anterior–posterior distribution with age. Increasing HI was associated with attenuation or reversal of this spatial organization. Global SWA showed no meaningful association with SDB severity. In contrast, regional frontal and posterior decay parameters were strongly associated with HI (adjusted R² = 0.53; p < 1 × 10⁻⁶) but not OAI (adjusted R² = 0.05; p = .95). The frontal-to-posterior decay-rate ratio showed the strongest association with HI β = 4.15; 95% CI, 3.17- 5.13; p < 1 × 10⁻¹⁰; adjusted R² = 0.55.

**Conclusions and Relevance:** Pediatric SDB was associated with regional disruption of slow-wave sleep homeostasis rather than global loss of deep sleep. These alterations affected both the spatial organization and temporal dynamics of SWA during a period of active cortical maturation and were not captured by conventional sleep metrics. Regional SWA dynamics may provide a developmentally sensitive marker of physiological disease burden in children with SDB.

**KEY POINTS:** *Question:* Does pediatric sleep-disordered breathing disrupt the regional organization and homeostatic dynamics of slow-wave activity during development in ways that are not captured by conventional sleep metrics?

*Findings:* In this cross-sectional study of 62 children across the spectrum of sleep-disordered breathing, hypopnea burden was associated with altered regional organization and overnight dissipation of NREM slow-wave activity (SWA) despite preserved global SWA. A frontal-to- posterior SWA decay-rate ratio was strongly associated with hypopnea severity, whereas global SWA was not.

*Meaning:* Pediatric sleep-disordered breathing may disrupt sleep physiology in a regional, developmentally meaningful manner not captured by conventional polysomnography, suggesting a potential physiological marker of disease burden beyond event counts.

## INTRODUCTION

Sleep-disordered breathing (SDB) affects approximately 1% to 5% of children and is among the most common reasons for referral to pediatric otolaryngology and sleep medicine.^1,2^ Despite its frequency, the physiological mechanisms linking pediatric SDB to daytime morbidity remain poorly understood. Children across the spectrum of SDB severity, including those with mild disorder, show impairments in attention, executive function, and behavior, yet the apnea-hypopnea index (AHI), the standard clinical severity metric, does not reliably predict which children will be affected.^3–10^ Standard sleep macrostructural measures, including total sleep time, sleep stage distribution, and arousal index, are similarly inconsistent predictors of morbidity.^11–14^ This mismatch has practical consequences: children with mild or predominantly hypopneic disease may appear physiologically preserved despite neurobehavioral dysfunction. Identifying brain-based markers of sleep disruption is therefore essential for moving beyond event counts toward a developmentally informed account of disorder burden.

One reason conventional polysomnography may miss relevant sleep disruption is that it treats sleep as spatially uniform. Sleep, however, is a local cortical process.^15^ Slow-wave activity (SWA; power in the 0.5-4 Hz band), a marker of NREM sleep intensity and homeostatic recovery, varies across the scalp as a function of local cortical state.^16–19^ This regional organization is especially important in childhood. In healthy development, SWA is maximal over posterior regions in early childhood and shifts anteriorly across adolescence, paralleling maturational changes in cortical structure and function.^20–25^ Thus, SWA topography is not merely a map of sleep depth; it is a developmentally meaningful marker of where sleep need and cortical maturation are most pronounced.

This developmental organization has direct implications for pediatric SDB. Prior studies have largely assessed sleep using macrostructural measures or limited electroencephalogram (EEG) montages, approaches that may obscure regional disturbances in the developing brain.^11–14^ A child may therefore have apparently preserved sleep architecture and normal global SWA while still showing disruption in cortical regions where SWA is developmentally prominent. Such a disturbance could help explain why standard polysomnography metrics incompletely capture the physiological burden of pediatric SDB.^3–14^

In the present study, we used high-density EEG (hdEEG) polysomnography in children spanning the spectrum of SDB severity to determine whether SDB is associated with regional disruption of SWA organization and overnight SWA dynamics. We tested whether hypopnea burden alters the expected age-related anterior-posterior organization of SWA, whether SDB is associated with opposing frontal and posterior SWA decay dynamics, and whether hypopnea index (HI) is more closely associated with these alterations than apnea-based indices.

## METHODS

### Study Design and Participants

This cross-sectional study included children aged 4 to 12 years who underwent overnight laboratory polysomnography with hdEEG. The study was designed to test the a priori hypothesis that pediatric sleep-disordered breathing is associated with altered regional organization of NREM slow-wave activity. Participants were recruited from clinical referral pathways and the community to sample the full continuum of respiratory event burden. Although community recruitment targeted children without habitual snoring or known sleep-disordered breathing, polysomnography identified measurable respiratory event burden in many community-recruited participants; conversely, some clinically referred children did not meet polysomnographic criteria for obstructive sleep apnea (OSA). The parent study was initially powered for categorical comparisons between children with minimal respiratory event burden and children with OSA, but because respiratory burden was continuously distributed across the recruited sample, the final analytic strategy modeled respiratory indices continuously in the full eligible sample rather than relying on recruitment sources or binary diagnostic categories.

The final analytic sample included 62 participants with usable overnight hdEEG data, valid respiratory scoring, and complete demographic data for planned models. Detailed inclusion and exclusion criteria, recruitment procedures, and reasons for exclusion are provided in eMethods 1 and eMethods 2.

### Ethics Statement

The study protocol was approved by the University of Wisconsin–Madison Institutional Review Board (protocol 2017-0681). Written informed consent was obtained from a parent or legal guardian for all participants, and child assent was obtained according to age and institutional requirements.

### Polysomography Acquisition, Sleep Staging, and Respiratory Scoring

All-night polysomnography (PSG) was performed in the laboratory. High-density EEG was recorded using a 256-channel HydroCel Geodesic Sensor Net connected to Compumedics Neuvo amplifiers. Signals were acquired at 500 Hz and referenced to vertex during acquisition. Sleep stages were scored in 30-second epochs according to American Academy of Sleep Medicine (AASM) criteria. Apneas and hypopneas were scored according to AASM criteria, and respiratory indices were computed as events per hour of sleep.^26^ The apnea-hypopnea index was defined as apneas plus hypopneas per hour of sleep; the obstructive apnea index as obstructive apneas per hour of sleep; and the hypopnea index as hypopneas per hour of sleep.

### EEG Processing and SWA Quantification

EEG processing was performed using EEGLAB and custom MATLAB scripts.^27^ Sleep scoring and arousal annotations were imported into the EEG structure, and non-EEG scoring channels were removed before EEG preprocessing. Data were high-pass filtered at .5 Hz, low-pass filtered at 40 Hz, and resampled to 200 Hz. Narrowband noise was then attenuated using Zapline-plus, targeting 10-, 20-, and 30-Hz components with a detection window size of 12.^28,29^ Bad channels and artifact-contaminated epochs were identified using visual inspection and semiautomated artifact detection. Channels with poor signal, along with all face and neck electrodes, were removed. Excluded scalp channels were replaced using spherical spline interpolation, and data were re-referenced to the average of retained channels.

Power spectral density was computed using fast Fourier transform in 6-second artifact-free NREM epochs. Slow-wave activity was defined as power in the 0.5- to 4-Hz range and was averaged across all-night N2 and N3 sleep unless otherwise specified. Full preprocessing procedures are provided in eMethods 2.

### Regional SWA and Frontal/Occipital Ratio

Frontal and occipital channel clusters were defined a priori based on the developmental SWA topography described by Kurth et al.^20,21^ The frontal/occipital SWA ratio was calculated as mean SWA across frontal channels divided by mean SWA across occipital channels. Detailed channel definitions are provided in eMethods 6 and eFigure 1.

**Figure 1.**
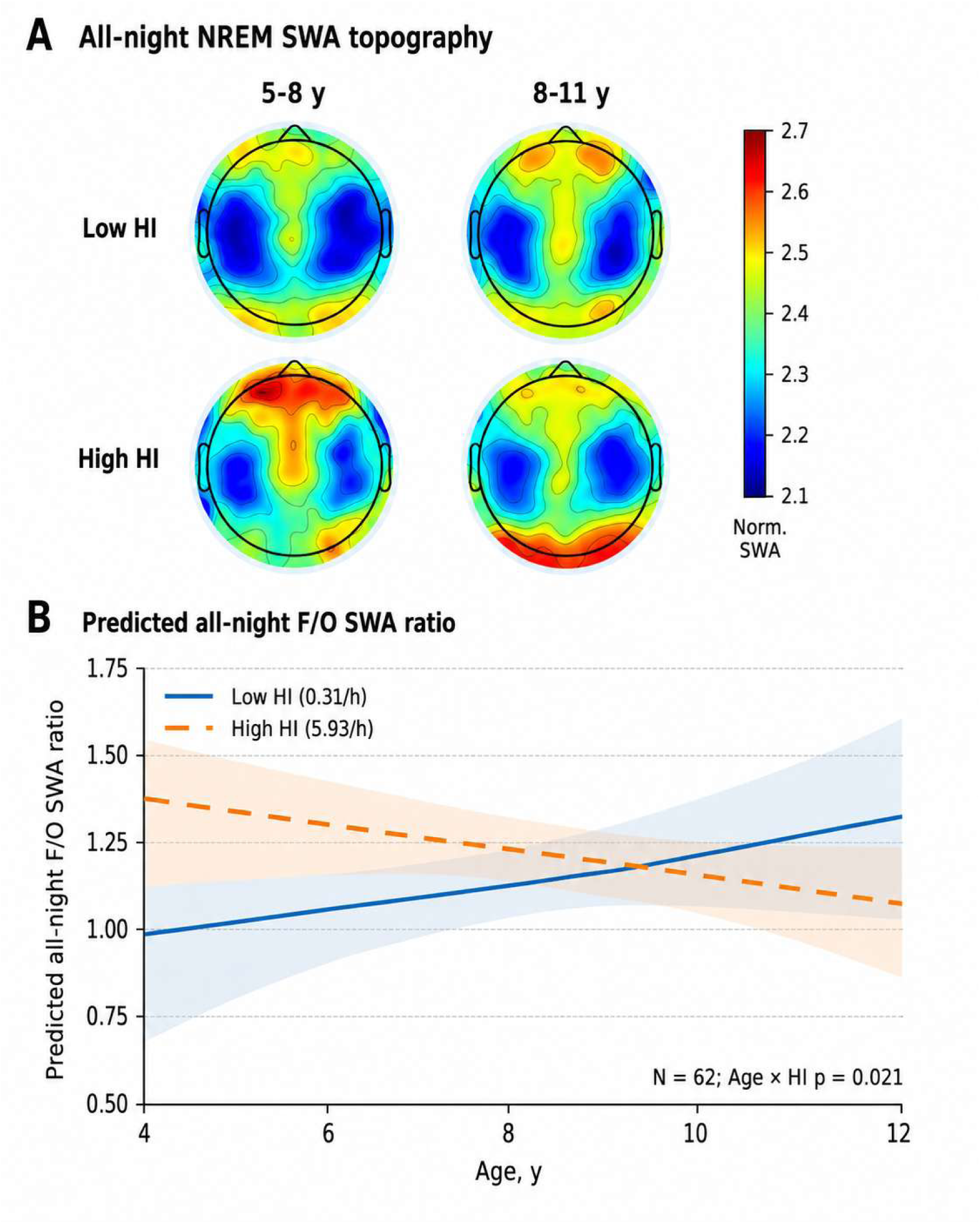
Regional organization of all-night NREM slow-wave activity by age and hypopnea burden. Panel A shows all-night NREM slow-wave activity (SWA; 0.5–4 Hz) topography in descriptive subgroups selected to visualize age- and HI-related differences in regional SWA organization. Maps show normalized SWA values displayed on a common color scale to emphasize spatial organization across the scalp rather than absolute group differences in SWA amplitude. The panel includes a subsample of the analytic cohort: younger/low HI, age 5–8 years and HI <1 event/h (n = 10); younger/high HI, age 5–8 years and HI ≥3.75 events/h (n = 9); older/low HI, age 8–11 years and HI <1 event/h (n = 12); and older/high HI, age 8–11 years and HI ≥3.75 events/h (n = 7). High HI was defined as the upper quartile of HI values in the analytic sample. Panel A was generated for visualization only and was not statistically compared. Panel B shows predicted all-night frontal/occipital SWA ratio across age from the regression model including HI, Age, Sex, and the Age × HI interaction in the full analytic sample (N = 62). Lines show model-predicted values at representative low and high HI values from the analytic sample: low HI, 0.31 events/h; high HI, 5.93 events/h. Shaded bands indicate 95% confidence intervals. The positive slope at low HI reflects the expected age-related increase in frontal relative to occipital SWA, whereas the negative slope at high HI indicates attenuation of this developmental pattern with greater hypopnea burden.

For descriptive visualization of regional SWA topography presented in Figure 1A, mean all-night NREM SWA was plotted in four subgroups defined by age and hypopnea burden. Younger children were defined as age 5 to 8 years and older children as age 8 to 11 years. Low HI was defined a priori as HI <1 event/h, and high HI was defined as the upper quartile of HI values in the analytic sample, corresponding to HI ≥3.75 events/h. Maps were normalized and displayed on a common color scale to emphasize spatial organization across the scalp rather than absolute differences in SWA amplitude. These subgroups were used only for visualization of SWA topography and were not used for inferential testing. Figure 1 legend and eMethods 2 contain subgroup sample sizes.

### Exponential Decay Modeling

Overnight SWA dynamics were modeled separately in frontal and posterior regions using an exponential decay function:

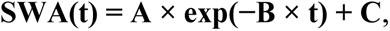

where **A + C** represents initial amplitude, **B** represents the overnight decay rate, and **C** represents the asymptotic SWA level. To summarize regional dissociation in SWA dissipation, we calculated frontal-to-posterior decay-rate ratios as the z-scored log difference between frontal and posterior parameters:

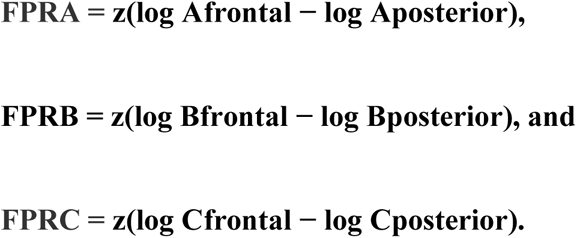

Full model-fitting procedures and model-selection details are provided in eMethods 7.

### Statistical Analysis

To test developmental SWA topography, all-night frontal/occipital SWA ratio was modeled as a function of HI, Age, Sex, and the Age × HI interaction. Figure 1A was descriptive and was not used as the primary statistical test of the study hypothesis. Predicted values in Figure 1B were generated from the full-sample regression model. To evaluate whether global or regional SWA metrics were associated with respiratory severity, separate regression models tested associations of global SWA and regional SWA decay parameters with HI, AHI, and OAI, adjusting for age and sex. Model performance was evaluated using adjusted R² and F tests. Partial regression plots based on the Frisch-Waugh-Lovell theorem were used for scatter plot visualizations so that displayed associations corresponded to adjusted model estimates. Analyses were conducted in MATLAB 2023b. The significance threshold was α = .05, two-tailed.

## RESULTS

### Participant Characteristics

Sixty-two children aged 4 to 12 years were included in the final analysis (mean [SD] age, 8.17 [1.78] years; 28 female [45%]). AHI ranged from 0.5 to 36.2 events/h (mean [SD], 4.6 [6.0]), and HI ranged from 0.1 to 34.1 events/h (mean [SD], 3.0 [5.3]). OAI was low and restricted in range (mean [SD], 0.5 [1.1]; range, 0.0-8.0 events/h). Sleep macrostructure was largely preserved across the sample (Table 1).

**Table 1.**
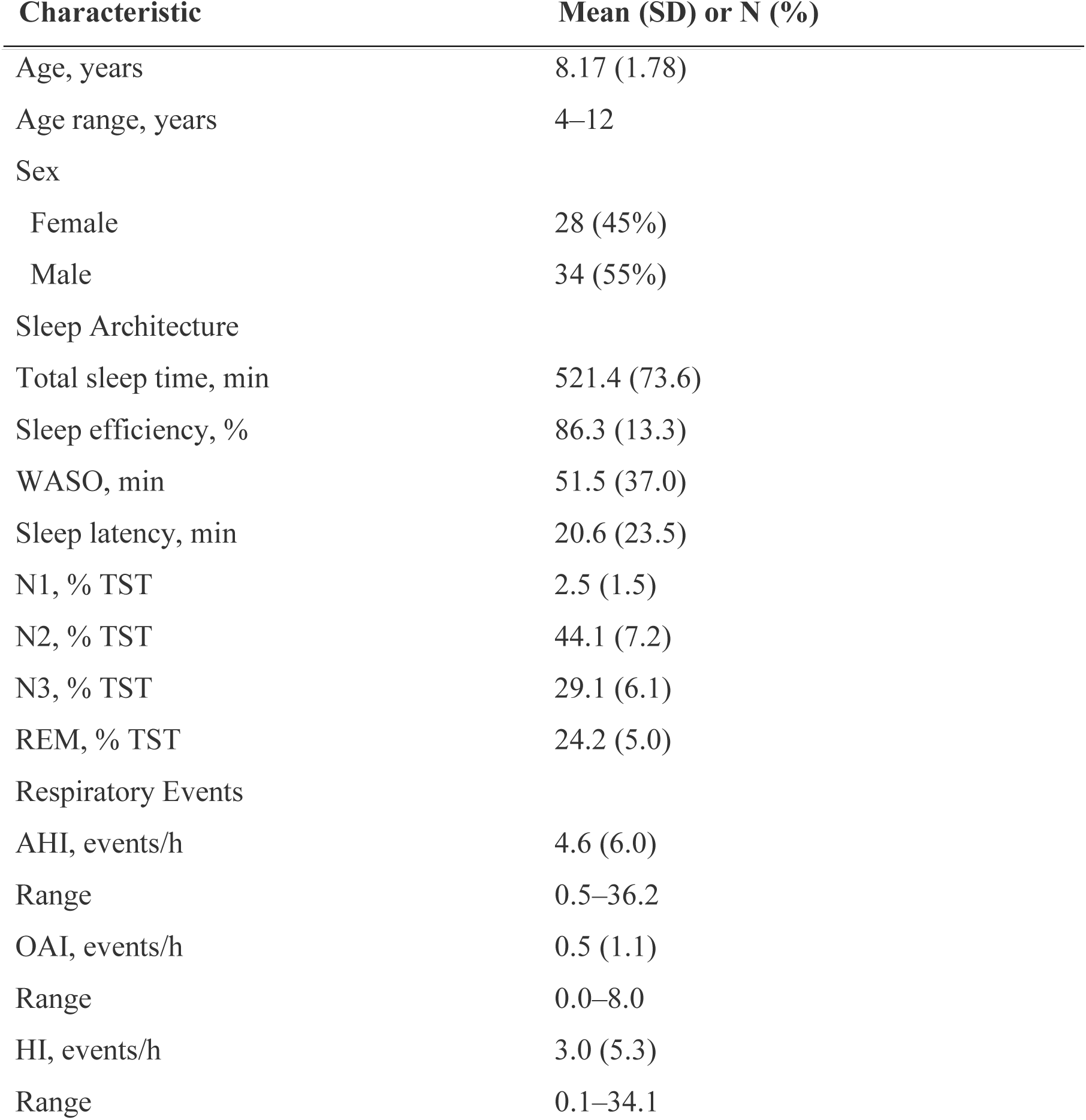
Participant Characteristics.

| Characteristic | Mean (SD) or N (%) |
| --- | --- |
| Age, years | 8.17 (1.78) |
| Age range, years | 4–12 |
| Sex |  |
| Female | 28 (45%) |
| Male | 34 (55%) |
| Sleep Architecture |  |
| Total sleep time, min | 521.4 (73.6) |
| Sleep efficiency, % | 86.3 (13.3) |
| WASO, min | 51.5 (37.0) |
| Sleep latency, min | 20.6 (23.5) |
| N1, % TST | 2.5 (1.5) |
| N2, % TST | 44.1 (7.2) |
| N3, % TST | 29.1 (6.1) |
| REM, % TST | 24.2 (5.0) |
| Respiratory Events |  |
| AHI, events/h | 4.6 (6.0) |
| Range | 0.5–36.2 |
| OAI, events/h | 0.5 (1.1) |
| Range | 0.0–8.0 |
| HI, events/h | 3.0 (5.3) |
| Range | 0.1–34.1 |

### Hypopnea Index Is Associated with Altered Developmental SWA Topography

Normalized all-night NREM SWA maps were generated to visualize the regional organization of SWA across age and hypopnea burden. Among children with lower hypopnea burden, SWA showed the expected developmental anterior-posterior organization, with relatively greater posterior SWA in younger children and a more balanced or anteriorly shifted distribution in older children (Figure 1A). This expected developmental pattern was attenuated among children with a higher hypopnea burden. In the full analytic sample (N = 62), the age- and sex-adjusted model of all-night frontal/occipital SWA ratio showed that HI was positively associated with the regional SWA ratio (β = 0.144; 95% CI, 0.017 to 0.271; p = .027), and the Age × HI interaction was significant (β = −0.015; 95% CI, −0.028 to −0.001; p = .036), indicating that increasing hypopnea burden weakened the expected age-related increase in frontal relative to occipital SWA (Figure 1B). Together, these findings indicate that HI was associated with altered developmental organization of NREM SWA rather than a simple global reduction in SWA.

### Global and Regional SWA Metrics Show Divergent Associations with Hypopnea Burden

Global SWA was not associated with HI in age- and sex-adjusted models (β = −0.01; 95% CI, −1.46 to 1.45; p = .99; adjusted R² = −0.04; Figure 2A), indicating that whole-scalp SWA summaries did not capture the hypopnea-related physiological signal observed in regional analyses. In contrast, the frontal/posterior SWA decay-rate ratio was strongly associated with HI (β = 4.15; 95% CI, 3.17 to 5.13; p < 1 × 10⁻¹⁰ ; adjusted R² = 0.55; Figure 2B). Thus, the association with hypopnea burden was not evident in overall SWA power but emerged when the regional organization of SWA dynamics was considered.

**Figure 2.**
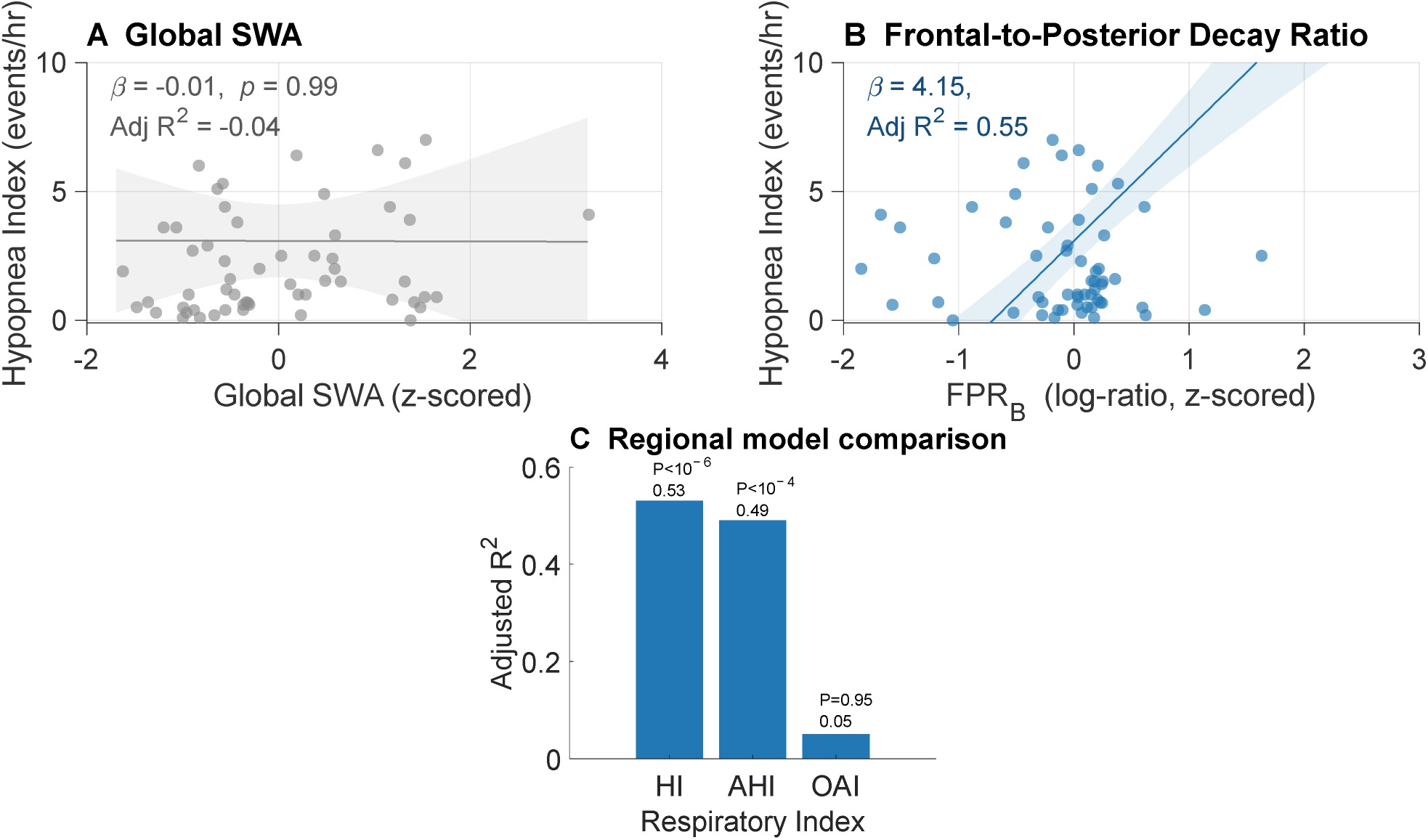
Global and regional SWA metrics show divergent associations with respiratory indices. Panel A shows the partial regression of global SWA on hypopnea index, showing no meaningful association. Panel B shows the partial regression of the frontal-to-posterior SWA decay-rate ratio on hypopnea index. Panel C compares adjusted model performance for regional SWA decay parameters across hypopnea index, apnea-hypopnea index, and obstructive apnea index. Scatter plots display partial residuals consistent with the adjusted regression models. SWA indicates slow-wave activity; HI, hypopnea index; AHI, apnea-hypopnea index; OAI, obstructive apnea index; FPRB, frontal-to-posterior decay-rate ratio.

Model comparisons further supported the stronger association between regional SWA dynamics and hypopnea-based respiratory indices. Frontal and posterior SWA decay parameters were strongly associated with HI (adjusted R² = 0.53; p < 1 × 10^−6^) and with AHI (adjusted R² = 0.49; p < 1 × 10^−4^), but not with OAI alone (adjusted R² = 0.05; p = .95; Figure 2C). This pattern suggests that the regional SWA signal was most closely linked to hypopnea burden rather than obstructive apneas alone.

### Hypopnea Burden Is Associated with Divergent Frontal and Posterior SWA Decay

SWA decay was modeled separately in frontal and posterior regions to characterize the dynamics underlying the regional association with hypopnea burden. In both regions, SWA declined across the night, but the trajectory of decline differed as a function of HI (Figure 3A and 3B). In children with lower HI, frontal and posterior SWA followed relatively parallel dissipation curves across the night. In children with higher HI, the two regions diverged: frontal SWA dissipated more rapidly, whereas posterior SWA remained elevated, producing an increasing anterior-posterior imbalance across the sleep period.

**Figure 3.**
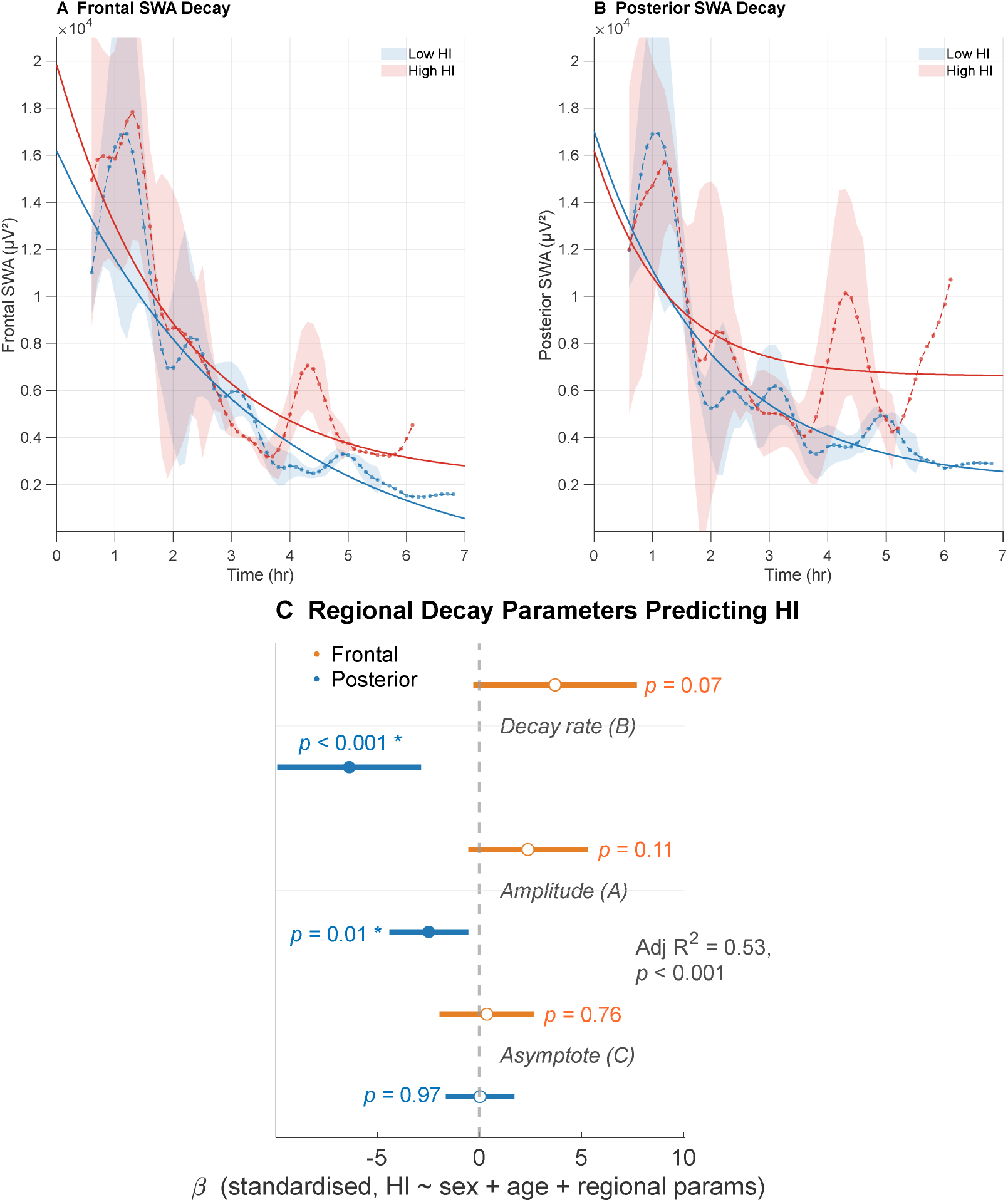
Regional dissociation of slow-wave homeostatic dynamics. Panel A shows representative frontal SWA decay curves for children with lower and higher HI, illustrating faster frontal decay with increasing hypopnea burden. Panel B shows representative posterior SWA decay curves for children with lower and higher HI, illustrating slower posterior decay with increasing hypopnea burden. Panel C shows regression coefficients (β ± SE) from the full regional model evaluating associations between frontal and posterior SWA decay parameters and HI. The full regional model was strongly associated with HI (adjusted R² = 0.53; p < 1 × 10⁻⁶). SWA indicates slow-wave activity; HI, hypopnea index.

In adjusted models controlling for age and sex, HI was associated with decay-rate and amplitude parameters, but not with asymptotic SWA level. Posterior decay rate was negatively associated with HI (β = −6.38; 95% CI, −9.90 to −2.86; p < .001), indicating slower overnight SWA dissipation with increasing HI. Frontal decay rate showed an association in the opposite direction (β = 3.70; 95% CI, −0.31 to 7.71; p = .07). Posterior amplitude was also negatively associated with HI (β = −2.48; 95% CI, −4.42 to −0.54; p = .01), whereas frontal amplitude was not (β = 2.37; 95% CI, −0.55 to 5.29; p = .11). Asymptotic SWA was unrelated to HI in both regions (frontal: β = 0.36; 95% CI, −1.96 to 2.67; p = .76; posterior: β = 0.03; 95% CI −1.65 to 1.70; p = .97; Figure 3C). The opposing directions of the frontal and posterior decay-rate associations indicate a regional dissociation of overnight SWA dynamics rather than a uniform reduction in sleep intensity.

### A Frontal-to-Posterior Decay Ratio Summarizes Regional SWA Imbalance

To determine whether this regional imbalance could be summarized parsimoniously, a reduced model was constructed using the three frontal-to-posterior ratio parameters: FPRA, FPRB, and FPRC. FPRB showed the strongest association with HI (β = 4.15; 95% CI, 3.17 to 5.13; p < 1 × 10^−10^), whereas FPRA and FPRC were not significant. This reduced model achieved adjusted R² = 0.55 (p < 1 × 10^−9^), comparable to the full regional model but with fewer parameters. These results support FPRB as a compact summary measure of the regional SWA imbalance associated with HI (Figure 2B).

## DISCUSSION

In this cross-sectional study of 62 children across the spectrum of SDB severity, hypopnea index was associated with altered regional slow-wave activity dynamics despite preserved global SWA. Three findings support this conclusion: higher HI disrupted the expected age-related anterior–posterior organization of SWA; frontal and posterior SWA decay showed opposing associations with hypopnea burden; and these effects were not captured by global SWA or obstructive apnea burden alone. Together, these findings suggest that pediatric SDB may disturb sleep physiology at a regional level that is invisible to conventional PSG summaries.

The developmental context is central. In childhood, SWA is not simply a measure of sleep depth; its scalp distribution changes systematically with cortical maturation. In healthy children, SWA is maximal posteriorly in early childhood and shifts anteriorly across development, paralleling maturational changes in cortical structure and function.^20–25^ The association between hypopnea burden and disruption of this age-related SWA gradient suggests that SDB may interfere with sleep physiology in cortical systems undergoing active maturation. This interpretation is consistent with pediatric neuroimaging studies showing that SDB is associated with alterations in regions supporting attention, executive control, and behavioral regulation, and extends that literature by identifying regional SWA disruption as a candidate sleep-physiological pathway linking nocturnal breathing disturbance to altered brain development.^31–36^

The opposing pattern of frontal and posterior SWA decay provides the clearest physiological insight. Under typical conditions, SWA declines across the night as sleep pressure dissipates.^16,17^ Here, higher hypopnea burden was associated with slower posterior decay and faster frontal decay, suggesting that NREM sleep recovery was not globally reduced but regionally uncoordinated. One plausible interpretation is that repeated partial obstruction perturbs NREM sleep in ways that disproportionately affect posterior cortical regions, which normally show prominent SWA in this age range. This distinction matters clinically because AHI combines heterogeneous events into a single frequency count. In this sample, the regional SWA signal was linked more closely to hypopneas than to obstructive apneas, suggesting that repeated partial obstruction may be a physiologically meaningful exposure for the developing brain.

These findings offer a possible explanation for a longstanding paradox in pediatric sleep medicine: children with SDB often show attentional and behavioral morbidity that is poorly predicted by AHI or standard sleep architecture.^3–10,34,37–40^ These data suggest a mismatch between what is typically measured in clinical PSG and what may be physiologically disrupted in the sleeping brain.^11–14^ Global SWA was unrelated to HI, whereas HI and regional SWA decay dynamics were tightly linked. The stronger association with HI than OAI should be interpreted cautiously because apnea severity was restricted in this sample. Nevertheless, the pattern raises the possibility that hypopneas, although individually less dramatic than complete obstructions, may cumulatively perturb NREM sleep in ways that are relevant to brain development. Prospective studies should test whether regional SWA dynamics predict neurobehavioral morbidity or treatment response beyond standard respiratory indices.

Several limitations should be noted. The cross-sectional design precludes causal inference; it is not possible to determine whether the observed SWA disruption is a consequence of hypopnea burden, a correlate of shared underlying factors, or a contributor to respiratory disturbance during sleep. The sample size of 62 limits precision of effect estimates and generalizability across the full range of pediatric age and SDB endophenotypes. All recordings were performed on a single night in a laboratory setting, which may not reflect habitual sleep. The restricted range of OAI limits interpretation of the null apnea finding. The ability of FPRB to serve as a generalizable physiological marker requires validation in independent samples before clinical application can be considered. We also did not establish whether the observed regional SWA alterations mediate neurobehavioral impairment; linking these physiological measures to attentional and behavioral outcomes will be essential in future work.

Pediatric SDB is associated with a regional alteration of both the spatial organization and temporal dynamics of SWA during a period of active cortical maturation. This disturbance is not captured by conventional sleep metrics and was most closely associated with hypopnea burden rather than obstructive apnea burden. These findings suggest that the physiological impact of pediatric SDB has been underestimated, not because it is subtle, but because it has been measured at the wrong level of organization. Regional SWA dynamics may help identify children whose sleep physiology is disrupted despite modest conventional PSG findings.

## Data Availability

Deidentified participant-level analytic data underlying the results reported in this article, together with a data dictionary and analytic code, may be made available to qualified investigators upon reasonable request after publication. Requests should be directed to the corresponding author and will be reviewed by the study investigators and the University of Wisconsin-Madison according to institutional policies, participant consent, and applicable IRB and data use requirements. Data will be shared only for scientifically appropriate analyses and under a signed data use agreement. Raw hdEEG/PSG files will not be made publicly available because of the size and complexity of the recordings and the potential risk of participant re-identification in a pediatric clinical research sample.

## Acknowledgments

The authors would like to thank Poorang Nori from Wisconsin Sleep for his contribution with the PSG set-up.

## Conflict of Interest Disclosures

GGM reports being an employee of Sleep Number and a paid consultant for Apnimed. The work reported in this article is unrelated to these roles.

No other disclosures were reported.

## Funding/Support

This work was supported by the Eunice Kennedy Shriver National Institute of Child Health and Human Development of the National Institutes of Health under award R21HD092986.

## Role of the Funder/Sponsor

The funding organization had no role in the design and conduct of the study; collection, management, analysis, or interpretation of the data; preparation, review, or approval of the manuscript; or the decision to submit the manuscript for publication.

## Data Sharing

Deidentified participant-level analytic data underlying the results reported in this article, together with a data dictionary and analytic code, may be made available to qualified investigators upon reasonable request after publication. Requests should be directed to the corresponding author and will be reviewed by the study investigators and the University of Wisconsin–Madison according to institutional policies, participant consent, and applicable IRB and data use requirements. Data will be shared only for scientifically appropriate analyses and under a signed data use agreement. Raw hdEEG/PSG files will not be made publicly available because of the size and complexity of the recordings and the potential risk of participant re-identification in a pediatric clinical research sample.

## Author Contributions

Conceptualization: SGJ, CM, GGM

Data curation: BP, TT, ES, AM

Formal analysis: GGM, BP, TT

Funding acquisition: SGJ

Investigation: SGJ, CM, BP, ES, TKavailability

Methodology: SGJ, CM, ES, GGM, AMV, AM

Project administration: SGJ, ES, AMV, AM

Resources: SGJ, CM, TK

Software: GGM, BP, TT

Supervision: SGJ

Validation: SGJ, GGM, BP, TT

Visualization: BP, TT, GGM

Writing – original draft: SGJ, GGM

Writing – review & editing: SGJ, CM, BP, TT, ES, GGM, TK, ES, AM

Where:

AM = Annika Myers

AMV = Ana Maria Vascan

BP = Bethany Peterson

CM = Camilla Matthews

ES = Emma Strains

GGM = Gary Garcia Molina

SGJ = Stephanie G. Jones

TT = Tamara P. Taporoski

## Notes

### Competing Interest Statement

Author GGM reports being an employee of Sleep Number and a paid consultant for Apnimed. The work reported in this article is unrelated to these roles. No other disclosures were reported.

### Author Declarations

The study was approved by the University of Wisconsin-Madison Institutional Review Board (protocol 2017-0681); a parent or legal guardian provided written informed consent and children provided age-appropriate assent before any study procedures.

