## Supplemental Material for "Regional Disruption of Slow-Wave Sleep Homeostasis in Children with Sleep-Disordered Breathing"

### eMethods 1. Study Design and Setting

This single-site observational study examined sleep neurophysiology across the spectrum of pediatric sleep-disordered breathing (SDB). The parent protocol used an unblinded, cross-sectional design with an initial daytime assessment visit followed by an overnight laboratory polysomnography/high-density electroencephalography (PSG/hdEEG) visit. The present analysis used the baseline overnight PSG/hdEEG recording and did not include sleep-disruption nights, follow-up/post-surgical nights, or other optional procedures.

Study procedures were conducted at the University of Wisconsin–Madison, including the Wisconsin Sleep Laboratory and associated Wisconsin Institute for Sleep and Consciousness research facilities. Overnight recordings were scheduled according to the child's habitual bedtime whenever possible. Children arrived approximately 2 hours before bedtime to allow hdEEG net fitting and PSG sensor placement. Caregivers were permitted to remain overnight; caregiver's presence was required for younger children according to protocol and for sleep-disruption procedures, which are outside the scope of the present analysis.

#### Ethics statement.

The study protocol was approved by the University of Wisconsin–Madison Institutional Review Board (protocol 2017-0681). Written informed consent was obtained from a parent or legal guardian for all participants, and child assent was obtained according to age and institutional requirements. No research procedures occurred before consent/assent.

**eMethods 2. Participants**

Children aged 4 to 12 years were recruited from clinical referrals for suspected obstructive sleep apnea (OSA) and from the community to capture the full range of pediatric SDB severity. Clinical recruitment included children referred through UW pediatric otolaryngology/general pediatric pathways, including children being evaluated for suspected OSA or adenotonsillectomy. Community recruitment included school and public flyers, community advertising, word of mouth, pediatric clinic advertising, and university-wide recruitment emails.

Recruitment occurred from May 2018 through March 2020, when in-person study procedures were interrupted by COVID-19-related laboratory closure. During this period, 80 youth signed informed consent/assent; 72 completed study procedures before the interruption. The final analytic sample for the current SWA paper included 62 youth with usable overnight hdEEG, valid respiratory scoring, and complete age and sex data for adjusted models. Because both clinically referred and community-recruited children showed a continuous range of respiratory event burden, analyses used objectively scored respiratory indices rather than recruitment source or presumed diagnostic status as the primary exposure.

Study Size Rationale. The parent study was originally designed to recruit children with minimal respiratory event burden and children with obstructive sleep apnea, defined a priori as AHI <1 event/h and AHI >5 events/h, respectively. The planned sample size was 27 participants per group, inflated by approximately 10% to target 30 participants per group, providing 90% power to detect the expected between-group difference in regional NREM SWA. During recruitment, respiratory event burden did not separate cleanly by recruitment source; therefore, the present analysis included all participants with usable hdEEG, valid respiratory scoring, and complete age and sex data and modeled respiratory indices continuously.

Race and ethnicity are summarized for the full consented sample using parent/legal guardian report. Because participant-level race and ethnicity were not included in the analytic EEG dataset used for these models, race and ethnicity are not re-tabulated for the final analytic subset.

eTable 1. Sample characteristics

| Characteristic | Consented sample (N = 80) | Final analytic sample (N = 62) |
| --- | --- | --- |
| Study flow | | |
| Signed informed consent/assent | 80 (100.0) | — |
| Completed study procedures before COVID-19 interruption | 72 (90.0) | — |
| Included in final analytic sample | — | 62 (100.0) |
| Sex | | |

| | | |
|---|---|---|
| Female | 42 (52.5) | 28 (45.2) |
| Male | 38 (47.5) | 34 (54.8) |
| Race | | |
| White | 47 (58.8) | — |
| Asian | 7 (8.8) | — |
| Black or African American | 5 (6.3) | — |
| American Indian/Alaska Native | 1 (1.3) | — |
| More than one race | 2 (2.5) | — |
| Unknown/not reported | 18 (22.5) | — |
| Ethnicity | | |
| Hispanic or Latino | 12 (15.0) | — |
| Not Hispanic or Latino | 53 (66.3) | — |

| | | |
|---|---|---|
| Unknown/not reported | 15 (18.8) | — |
| Age, y, mean (SD); range | 8.0 (1.9); range, 3.7-11.8 | 8.2 (1.8); range, 4.1-11.9 |
| AHI, events/h, median (IQR); range | — | 2.9 (1.6-6.0); range, 0.5-36.2 |
| AHI, events/h, mean (SD) | — | 4.7 (6.0) |
| BMI percentile, median (IQR); range | — | 51.0 (30.5-66.0); range, 11.0-91.0 |
| BMI percentile ≥5th and 85th percentile | — | 54/59 (91.5) |

Values are No. (%) unless otherwise indicated. Abbreviations: AHI, apnea-hypopnea index; BMI, body mass index; IQR, interquartile range. Race and ethnicity were collected separately; Hispanic or Latino ethnicity may be reported with any race category.

**Inclusion criteria.**

Core inclusion criteria included child age in the preadolescent study range, presence of a caregiver/guardian able to provide informed consent and history reporting, English language ability sufficient to complete study procedures, and ability to tolerate the EEG/PSG setup and remain still for study procedures. Children in the clinically referred group had suspected OSA or related clinical referral history; community-recruited children were recruited as comparison participants or children with snoring symptoms depending on study version.

**Exclusion criteria.**

Exclusion criteria included history of significant head trauma, unstable medical illness, current psychotropic medication use, contraindications to MRI for participants completing optional MRI procedures, claustrophobia for MRI procedures, possible pregnancy when applicable by self-report, intellectual disability or significantly below-age-expected cognitive functioning, assessed using the Q-interactive WISC-V or WPPSI-IV , as age appropriate.

**eMethods 3. Study Procedures and Measures**

Visit 1 consisted of consent/assent followed by parent/caregiver and child questionnaires, demographic and clinical history forms, standardized neurobehavioral measures, and distribution of wrist actigraphy when available. Parent/caregiver forms captured demographics, treatment history, medication use, medical history, developmental milestones, and family psychiatric history. Pubertal status was assessed using the Pubertal Development Scale and/or Tanner picture-based ratings when age-appropriate. Handedness was assessed using a handedness questionnaire or behavioral handedness index.

**Sleep questionnaires and actigraphy.**

Parent-reported sleep history and sleep symptoms were assessed using standardized pediatric sleep questionnaires, including the Children’s Sleep Habits Questionnaire and/or Sleep Disturbance Scale for Children depending on study version. Children were asked to wear wrist actigraphy for up to 7 days before the overnight visit when devices were available; actigraphy and/or sleep diaries were used to characterize habitual sleep timing and support scheduling of the overnight visit. These measures were used to characterize the parent protocol and may be used in future analyses, but they were not primary outcomes in the current SWA analysis unless explicitly listed as covariates.

**Neurobehavioral and symptom measures.**

The study protocol included cognitive, behavioral, and symptom measures designed to characterize the broader pediatric SDB phenotype. Measures included NIH Toolbox cognition and emotion batteries, Wechsler measures or selected WISC/WPPSI subtests, Vanderbilt ADHD Rating Scale, and related parent- or child-report symptom measures depending on study version and child age. The Tests of Variables of Attention (T.O.V.A), a computerized continuous performance task, was administered before and after the overnight sleep study. These measures were not primary outcomes for the present manuscript, which tested the a priori physiological hypothesis that SDB is associated with altered regional organization and overnight dynamics of NREM SWA. Behavioral and questionnaire outcomes will be reported in separate analyses focused on neurobehavioral morbidity and morning performance.

**Visit 2: Overnight PSG/hdEEG.**

The overnight visit lasted approximately 12 hours depending on the child's sleep-wake schedule. Children completed waking electroencephalogram (EEG) and computerized tasks before sleeping, followed by all-night polysomnography (PSG) with high-density EEG (hdEEG). After ad libitum sleep, morning neurobehavioral testing was repeated. The present manuscript uses the baseline overnight sleep recording only.

**eMethods 4. Polysomnography, Respiratory Scoring, and Sleep Variables**

All-night PSG was performed in the laboratory. The clinical PSG montage included electrooculogram, chin electromyogram, leg electromyogram, pulse oximetry, respiratory effort belts, and video monitoring as available. High-density EEG was recorded concurrently as described below. Sleep stages were visually scored in 30-second epochs according to American Academy of Sleep Medicine criteria. Respiratory events were scored according to American Academy of Sleep Medicine criteria, and clinical sleep-disordered breathing diagnoses/interpretations were reviewed by a board-certified sleep medicine physician.

Respiratory indices were computed as events per hour of sleep. Apnea-hypopnea index (AHI) was defined as apneas plus hypopneas per hour of sleep; obstructive apnea index (OAI) as obstructive apneas per hour of sleep; and hypopnea index (HI) as hypopneas per hour of sleep. Additional PSG variables included total sleep time, sleep efficiency, wake after sleep onset, sleep latency, percent of total sleep time spent in N1, N2, N3, and REM sleep, oxygen desaturation metrics, and arousal index when available.

The analytic sample included children across a continuous range of SDB severity. In the current manuscript, AHI ranged from 0.5 to 36.2 events/h, HI ranged from 0.1 to 34.1 events/h, and OAI ranged from 0.0 to 8.0 events/h. Median AHI in the final analytic sample was 2.9 events/h (IQR, 1.6-6.0), supporting analysis across the pediatric SDB severity spectrum. Primary inference modeled respiratory indices continuously rather than using recruitment source or binary clinical grouping. Respiratory indices were not transformed prior to analysis. AHI, HI, and OAI were modeled in their original units (events per hour of sleep) as continuous predictors in all regression models.

**eMethods 5. hdEEG Acquisition and Preprocessing**

Overnight hdEEG was recorded using a 256-channel HydroCel Geodesic Sensor Net (Electrical Geodesics Inc., Eugene, OR) adapted for overnight recording with Compumedics Neuvo amplifiers. Electrodes were filled with electrolyte gel, and additional gold electrodes were attached to the chin for electromyography. EEG signals were referenced to the vertex during acquisition and sampled at 500 Hz.

EEG data were processed offline in MATLAB using EEGLAB, Zapline-plus, and custom laboratory scripts. Raw EEG files were loaded with the appropriate EGI channel-location file. Sleep scoring and arousal annotations were imported or represented within the EEG structure, and non-EEG scoring channels were removed before EEG preprocessing. Data were high-pass filtered at 0.5 Hz, low-pass filtered at 40 Hz, and resampled to 200 Hz. Narrowband line-related noise was attenuated using Zapline-plus, targeting 10-, 20-, and 30-Hz components with a detection window size of 12.

Bad channels and artifact-contaminated epochs were identified using visual inspection and semiautomated artifact detection. Channels with poor signal and all face and neck electrodes were removed. Data were re-referenced to the average of retained channels and removed scalp channels were then replaced using spherical spline interpolation in EEGLAB, such that each subject's final channel set consisted of the same 172 channels nearest to the vertex.

**eMethods 6. Spectral Analysis and Regional SWA Metrics**

Power spectral density was computed for each retained EEG channel using fast Fourier transform applied to 6-second artifact-free NREM epochs. Slow-wave activity (SWA) was defined as power in the 0.5- to 4-Hz range and was averaged across artifact-free N2 and N3 sleep unless otherwise specified.

Global SWA was calculated as mean SWA across the retained scalp channel set. Frontal and occipital regions of interest were defined a priori based on developmental hdEEG literature and the expected posterior-to-anterior maturational shift in SWA topography. As in Kurth et al,[20] the frontal region stretched between FP1 and FP2 (channels 37 and 10, not included) and spread posteriorly as far as Fz (channel 21, included), and the occipital region was centered between O1 and O2 (channels 116 and 150, not included) and included the same number of channels as the frontal cluster. The frontal/occipital SWA ratio was calculated as mean SWA across frontal channels divided by mean SWA across occipital channels.

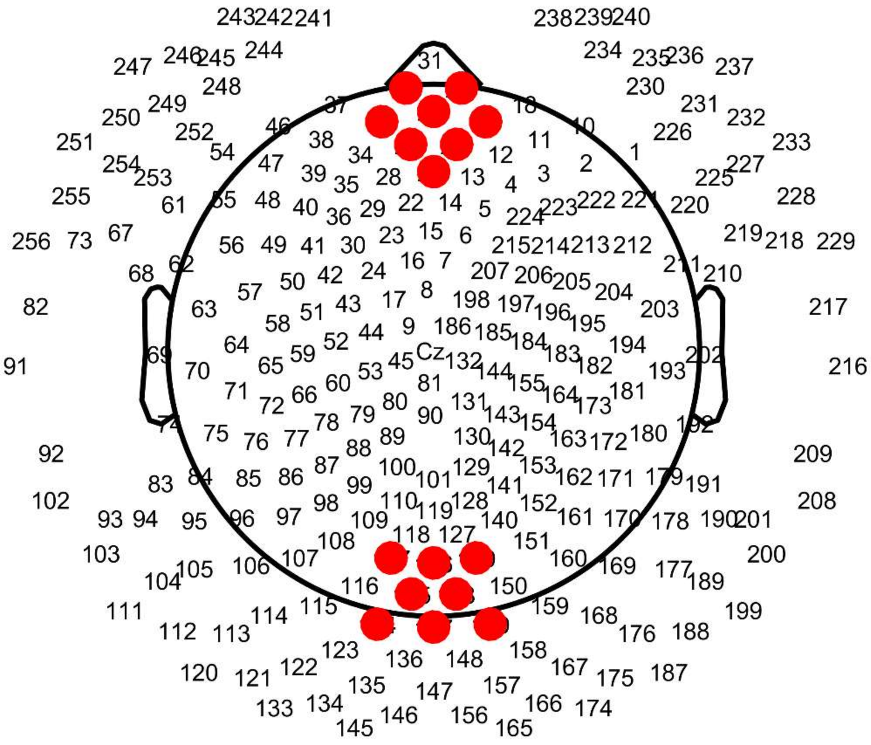


eFigure 1. Frontal and Occipital Region-of-Interest Channel Definitions.

Frontal (top) and occipital (bottom) regions of interest are shown in red on the 256-channel HydroCel Geodesic Sensor Net layout (top view). Channel numbers are labeled. The frontal region spans from FP1/FP2 (channels 37 and 10) posteriorly to Fz (channel 21). The occipital region is centered between O1/O2 (channels 116 and 150) and includes the same number of channels as the frontal cluster. Channel definitions follow Kurth et al.[20]

**Descriptive topographic visualization.**

For Figure 1A, mean all-night NREM SWA topography maps were normalized and four descriptive subgroups plotted on a common color scale to visualize age- and HI-related differences in regional SWA organization. Younger children were defined as age 5 to 8 years, and older children were defined as age 8 to 11 years. Low HI was defined a priori as HI <1 event/h. High HI was defined data-adaptively as the upper quartile of HI values in the analytic sample, corresponding to HI ≥3.75 events/h. This resulted in four descriptive subgroups: younger/low HI (n = 10), younger/high HI (n = 9), older/low HI (n = 12), and older/high HI (n = 7). These subgroups were used only for visualization of observed SWA topography and were not statistically compared. All inferential analyses used the full analytic sample and modeled respiratory indices continuously.

**eMethods 7. Exponential Decay Modeling of Overnight SWA Dynamics**

Overnight SWA dynamics were modeled using an exponential decay function:

$$SWA(t) = A \times \exp(-B \times t) + C$$

where A + C represents estimated initial SWA, B represents the decay rate, and C represents the asymptotic SWA level. Models were fit separately for frontal and posterior regions. Candidate model-fitting approaches varied in the temporal windowing and sampling strategy used to summarize overnight SWA dynamics. The final approach was selected based on model fit, stability of parameter estimation, and physiological interpretability of the resulting frontal and posterior decay curves.

Candidate model-fitting approaches varied in the temporal windowing (20, 30, or 60 minutes) and sampling strategy (uniform vs non-uniform) used to summarize overnight SWA dynamics, yielding six combinations evaluated against each respiratory index. The final approach was selected based on model fit as indexed by adjusted $R^2$ and overall model significance across outcomes. Non-uniform sampling consistently outperformed uniform sampling within each window duration, and 20-minute windows yielded superior or equivalent fit relative to longer windows under non-uniform sampling. The 20-minute non-uniform approach produced the strongest fit for both HI (adjusted $R^2 = 0.53$, $p < 1\times10^{-6}$) and AHI (adjusted $R^2 = 0.40$, $p < 1\times10^{-4}$), while OAI showed negligible explanatory variance across all approaches (adjusted $R^2$ ranging from −0.10 to −0.09, all $p > 0.82$), consistent with the restricted range of obstructive apnea events in this pediatric sample. Accordingly, 20-minute non-uniform sampling was adopted as the final windowing strategy. To summarize regional dissociation in SWA homeostasis, frontal-to-posterior ratio parameters were computed from the regional exponential decay estimates. The primary summary metric was the frontal-to-posterior decay-rate ratio:

**FPRB = z(log Bfrontal − log Bposterior)**

This metric captures the relative balance of frontal versus posterior SWA dissipation across the night, with higher values reflecting faster frontal decay relative to posterior decay. Analogous frontal-to-posterior ratios for amplitude and asymptote parameters were examined as secondary summary metrics.

**eMethods 8. Statistical Analysis**

The primary objective was to test whether objectively scored respiratory burden was associated with regional organization and overnight dynamics of NREM SWA after accounting for developmental effects. Age and sex were included as core covariates because pediatric SWA topography and SDB burden vary developmentally and by sex. Statistical significance was evaluated using two-sided tests with $\alpha = .05$ unless otherwise specified.

**Developmental SWA topography.**

All-night frontal/occipital SWA ratio was modeled in the full analytic sample using linear regression:

$$\text{F/O SWA}_i = \beta_0 + \beta_1\text{Age}_i + \beta_2\text{HI}_i + \beta_3\text{Sex}_i + \beta_4(\text{Age}_i \times \text{HI}_i) + \varepsilon_i,$$

where F/O $\text{SWA}_i$ is the all-night frontal/occipital SWA ratio for participant i, $\text{Age}_i$ is age in years, $\text{HI}_i$ is hypopnea index in events per hour of sleep, $\text{Sex}_i$ is participant sex, and $\varepsilon_i$ is the residual error term. The Age × HI coefficient tested whether hypopnea burden modified the expected age-related increase in frontal relative to occipital SWA. For Figure 1B, model-predicted F/O SWA ratio values were generated across the observed age range with HI fixed at the 10th and 90th percentiles of the analytic sample, corresponding to 0.31 and 5.93 events/h, respectively. Sex was held at the sample mean to generate sex-adjusted predicted values. Shaded bands represent 95% confidence intervals for the model-predicted mean values.

**Global and regional SWA associations.**

Regression models evaluated associations of global SWA, regional SWA decay parameters, and frontal-to-posterior ratio parameters with HI, AHI, and OAI, adjusting for age and sex. Model performance was evaluated using adjusted $R^2$ and F tests. Partial regression plots based on the Frisch-Waugh-Lovell theorem were used for scatter plot visualizations, so displayed associations corresponded to adjusted model estimates.

EEG preprocessing and primary analyses were performed using MATLAB (Version R2023b; MathWorks), EEGLAB (Version 2023.1; Delorme & Makeig, 2004), the Zapline-plus extension (Version 1.2.1; Klug & Kloosterman, 2022), and custom laboratory scripts.

**eMethods 9. Missing Data, Exclusions, and Bias Control**

Missingness was handled by analysis stream (see eFigure 2). Of 80 youth who signed informed consent/assent, 72 completed study procedures before COVID-19-related laboratory closure, and 62 were included in the current SWA analytic sample. Participant-level inclusion in the current SWA analysis required usable overnight hdEEG, valid respiratory scoring, and complete age and sex data for adjusted models. EEG exclusions distinguished participant-level exclusion from channel-level interpolation and epoch-level artifact rejection. Artifact-contaminated epochs were excluded from spectral estimates; channels with poor signal were removed and interpolated when participant-level data quality remained adequate. Participant-level data was considered inadequate if more than 35% of channels were interpolated (3 subjects); if too many channels were removed from the center of the head, leading to poor interpolation performance (1 subject); if more than 25% of the NREM recording was removed due to artifact contamination (1 subject); if the total recording time was less than 4 hours (3 subjects); if the net fit poorly (1 subject); or if prominent sweat artifact could not be removed from the recording (1 subject).

Bias was addressed by modeling respiratory indices continuously in the full analytic sample rather than relying on recruitment source or binary diagnostic labels. This approach reduced misclassification introduced by the observation that some community-recruited children showed PSG-defined respiratory event burden, and some clinically referred children did not meet PSG criteria for OSA. Age and sex were included in core models. Pubertal status, BMI/obesity, ADHD symptoms or medication status, sleep duration/efficiency, recruitment source, and sleep timing should be considered in sensitivity analyses if sufficiently complete and not collinear with the primary exposure.

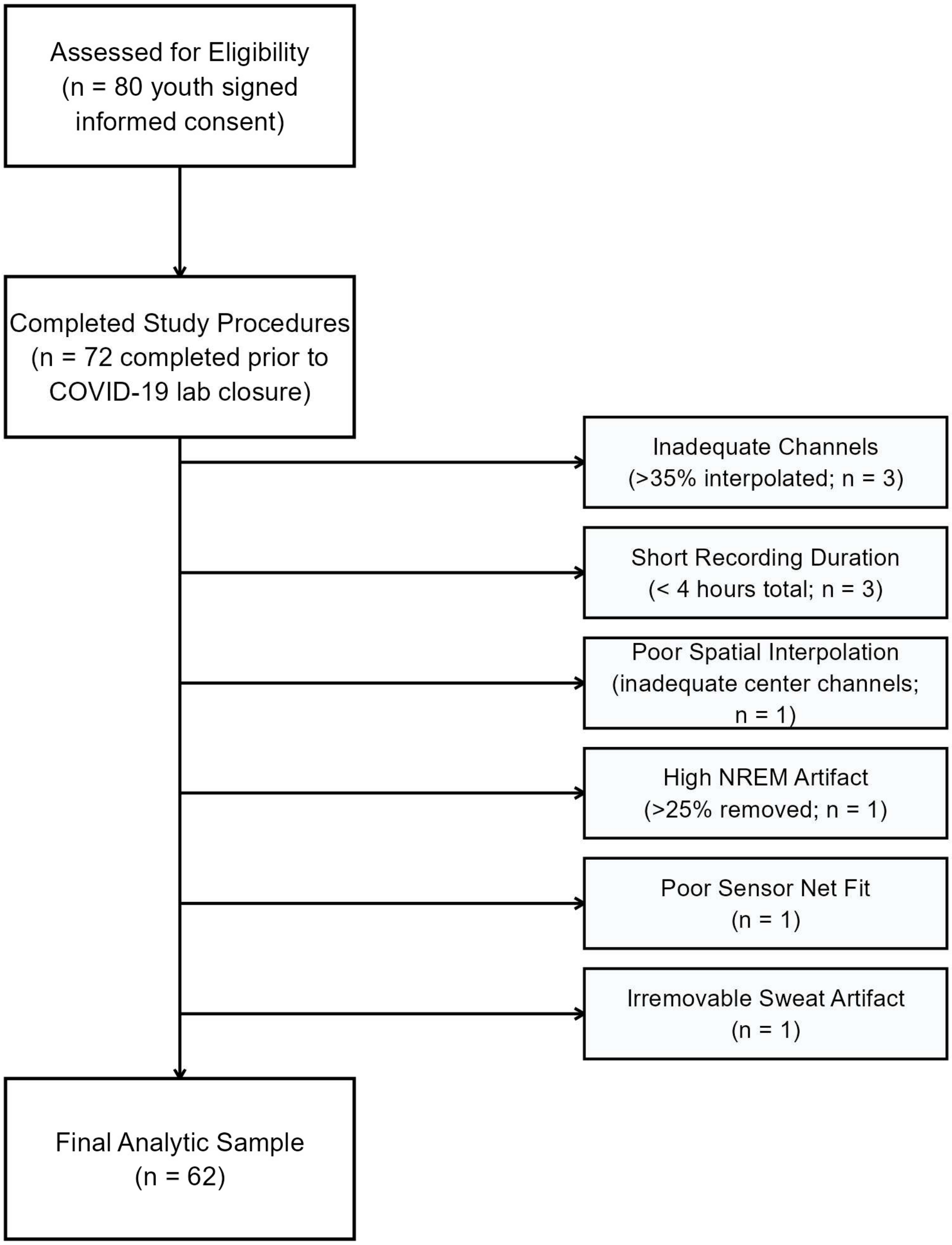


eFigure 2. Participant Flow Diagram.
